# Prevalence, Bacterial Aetiology and Antimicrobial Resistance Patterns of Neonatal Sepsis at Livingstone University Teaching Hospital, Zambia: A Retrospective Cross-Sectional Record Review, 2020–2025

**DOI:** 10.64898/2026.09.27.26364139

**Authors:** Lumamba Siaciti, Malala Siacit, Cornelius Simutanda, Nasson Nathan Tembo

## Abstract

**Background:** Neonatal sepsis remains a major cause of neonatal morbidity and mortality, particularly in low- and middle-income countries. Its management is increasingly complicated by antimicrobial resistance (AMR), while limited facility-specific data on causative bacterial pathogens and susceptibility patterns often necessitate empirical antibiotic treatment. This study assessed the prevalence of neonatal sepsis, bacterial pathogens and antimicrobial resistance patterns, and maternal, neonatal and clinical factors associated with neonatal sepsis among neonates admitted to the Neonatal Intensive Care Unit (NICU) at Livingstone University Teaching Hospital (LUTH), Zambia.

**Methods:** A retrospective cross-sectional record review was conducted among neonates admitted to the NICU at LUTH from 2020 to 2025. A total of 150 neonatal records were analysed. Neonatal, maternal, clinical, blood culture and antimicrobial susceptibility data were analysed using descriptive statistics, chi-square tests and logistic regression. Missing information was treated as missing and was not imputed; denominators were reported according to the records available for each analysis.

**Results:** Neonatal sepsis was identified in 52/150 neonates, giving a prevalence of 34.7% (95% CI: 27.5%–42.6%). Among the 52 sepsis cases, 20 had positive blood cultures, corresponding to a culture positivity rate of 38.5% (95% CI: 26.5%–52.0%); this represented 13.3% of the full study sample. Maternal fever during labour (aOR=6.822; 95% CI: 2.032–22.905; p=0.002) and caesarean delivery (aOR=2.245; 95% CI: 1.050–4.801; p=0.037) were independently associated with neonatal sepsis, while increasing birth weight was protective (aOR=0.891 per 100 g; 95% CI: 0.817–0.973; p=0.010). Escherichia coli was the most frequent isolate (5/20; 25.0%), followed by Klebsiella pneumoniae (4/20; 20.0%) and Staphylococcus aureus (4/20; 20.0%). Resistance was highest to ampicillin (15/19; 78.9%), followed by gentamicin and ceftriaxone (10/19; 52.6% each), while all 19 isolates tested against meropenem were susceptible.

**Conclusion:** The findings indicate that empirical management of neonatal sepsis at LUTH should be informed by local microbiological surveillance rather than reliance on historical first-line susceptibility assumptions. Routine blood culture and susceptibility testing, a regularly updated facility antibiogram, strengthened infection prevention and antimicrobial stewardship are required to improve treatment decisions and preserve effective antibiotics. Interpretation should consider the retrospective single-centre design, incomplete routine records and the small number of culture-positive isolates.

## Introduction

Neonatal sepsis is a major public health problem and an important cause of neonatal morbidity and mortality, especially in low- and middle-income countries (LMICs). Neonatal infections contribute substantially to preventable deaths, with the greatest burden occurring in sub-Saharan Africa and South Asia [1]. In Zambia, neonatal mortality remains high, and neonatal infections including sepsis contribute to early neonatal deaths [2]. Tertiary referral hospitals such as LUTH manage high-risk neonates, making sepsis a frequent clinical challenge in neonatal intensive care.

Antimicrobial therapy is central to neonatal sepsis management, but its effectiveness is increasingly threatened by antimicrobial resistance. Empirical regimens commonly include ampicillin and gentamicin, yet resistance to these agents is increasingly reported, particularly in LMICs [3-6]. Gram-negative organisms such as Klebsiella pneumoniae, Escherichia coli and Pseudomonas aeruginosa are important causes of neonatal bloodstream infection and may exhibit multidrug resistance [5,7].

Evidence from Zambia has demonstrated substantial resistance among neonatal bloodstream isolates to commonly used first-line antibiotics [6]. However, facility-specific surveillance at LUTH remains limited, and clinicians frequently rely on empirical treatment in the absence of updated local antibiograms. This limits evidence-based prescribing and antimicrobial stewardship. The present study therefore aimed to determine the prevalence of neonatal sepsis, identify bacterial pathogens and their antimicrobial resistance patterns, and assess maternal and neonatal factors associated with sepsis among NICU admissions at LUTH.

## Methods

### Study design and setting

This was a retrospective cross-sectional record review conducted in the NICU at Livingstone University Teaching Hospital, the principal tertiary referral hospital for Southern Province, Zambia. Existing NICU and laboratory records for neonates admitted from 2020 to 2025 were reviewed without prospective follow-up. The NICU has a bed capacity of 36 and admits approximately 130 neonates per month, including critically ill neonates with suspected sepsis.

### Study population and eligibility

The study population comprised neonates admitted to the NICU during the study period from 2020 to 2025. Records were eligible when relevant demographic, clinical and laboratory information was available. For analyses of bacterial aetiology and antimicrobial susceptibility, records with blood culture and antimicrobial susceptibility testing results were used. Contaminated or indeterminate culture results were excluded.

### Sample size and sampling

The minimum sample size for the prevalence component was estimated using the Cochran formula, assuming a prevalence of 9.5%, 5% precision and a 95% confidence level, yielding 133 records. To improve precision and allow for incomplete routine records, 150 eligible neonatal records were included in the final analysis. Total population sampling (total enumeration) for eligible neonatal admission records was used. Laboratory outcomes used smaller denominators because culture and antimicrobial susceptibility data were available only for subsets of records.

### Data collection and quality assurance

Data were abstracted from hospital records using a pretested structured electronic form developed in Google Forms. Variables included neonatal demographics, birth characteristics, maternal and obstetric factors, clinical outcomes, blood culture results and antimicrobial susceptibility patterns. Ten percent of extracted records were randomly audited by the principal investigator for accuracy and completeness. Personal identifiers were excluded and unique study identification numbers were used.

### Statistical analysis

Data were exported and analysed using SPSS version 25. Data cleaning included range, consistency and missing-value checks. Missing values were not imputed and analysis used available records for each variable. Categorical variables were summarised using frequencies and percentages, while continuous variables were summarised using means as available in the source analysis. Neonatal sepsis prevalence and culture positivity were reported with 95% confidence intervals. Pearson chi-square or Fisher’s exact tests were used as appropriate for categorical comparisons. Univariable logistic regression estimated crude odds ratios (ORs) with 95% CIs, and multivariable logistic regression estimated adjusted odds ratios (aORs). Statistical significance was set at p<0.05. Antimicrobial susceptibility results were summarised using the number and proportion of isolates classified as sensitive or resistant to each antibiotic tested. Denominators were stated explicitly for the overall sample (n=150), sepsis cases (n=52), culture-positive cases (n=20), and isolates with reported susceptibility testing (n=19).

### Ethical considerations

Ethical clearance was obtained from the Mulungushi University School of Medicine and Health Sciences Research Ethics Committee and the National Health Research Authority before commencement of the study. The requirement for informed consent was waived because the study involved secondary analysis of anonymized hospital data. Confidentiality was maintained through de-identification and restricted access to the dataset.

## Results

### Participant characteristics and prevalence of neonatal sepsis

A total of 150 neonates were included, of whom 52 had neonatal sepsis, giving a prevalence of 34.7% (95% CI: 27.5%–42.6%), while 98 (65.3%) did not. Among the 52 sepsis cases, 20 were culture-positive, giving a culture positivity rate of 38.5% (95% CI: 26.5%–52.0%); culture-positive cases represented 13.3% (95% CI: 8.8%–19.7%) of the full sample. Male neonates comprised 60.7% of the sample. Low birth weight was more common among neonates with sepsis (63.5%) than those without sepsis (39.8%; p=0.017). Maternal fever during labour was also more frequent among sepsis cases (19.2% vs 5.1%; p=0.014). Mortality was higher in the sepsis group (30.8% vs 12.2%; p=0.006), and mean length of hospital stay was longer (7.77 vs 4.76 days; p<0.001).

**Table 1:**
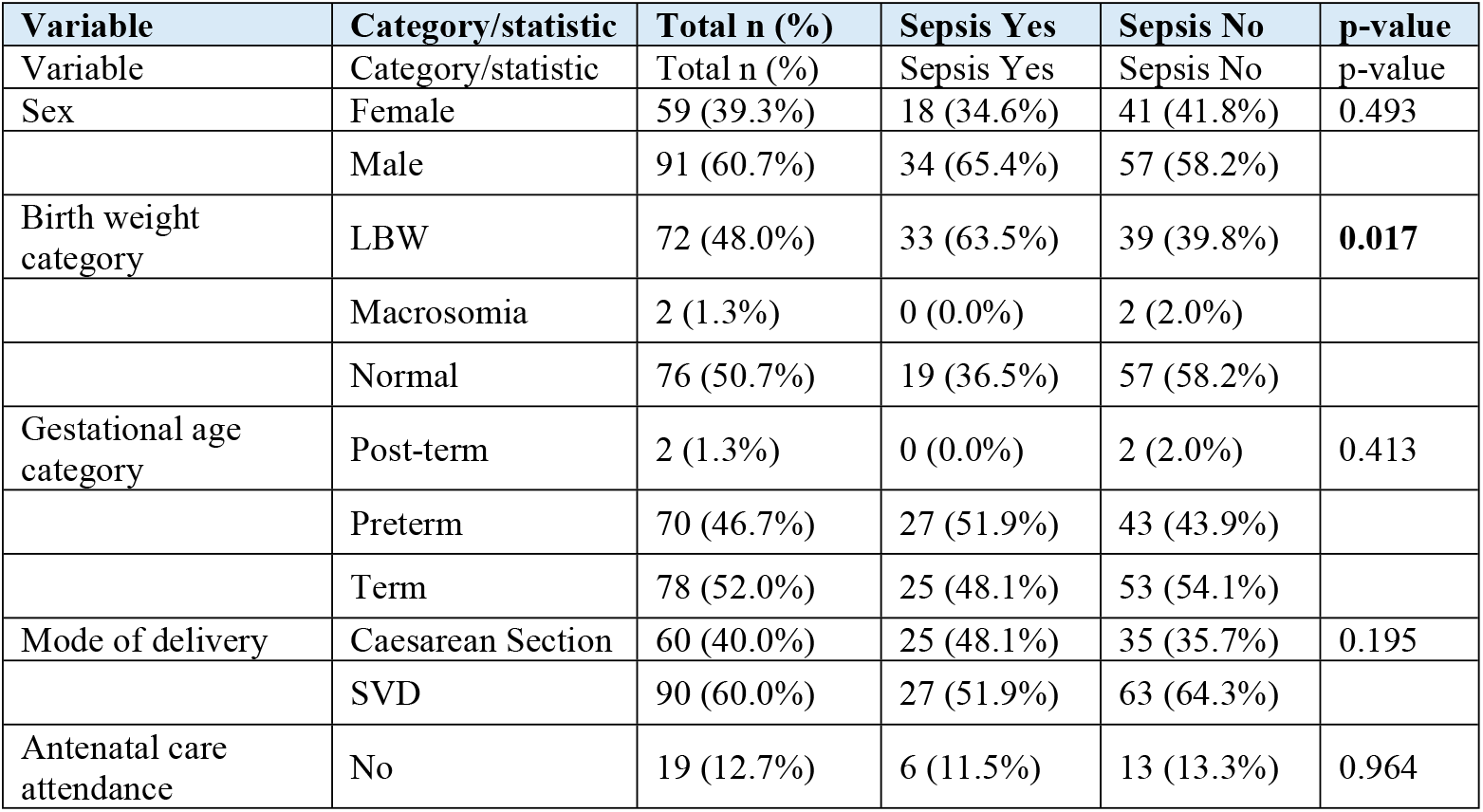
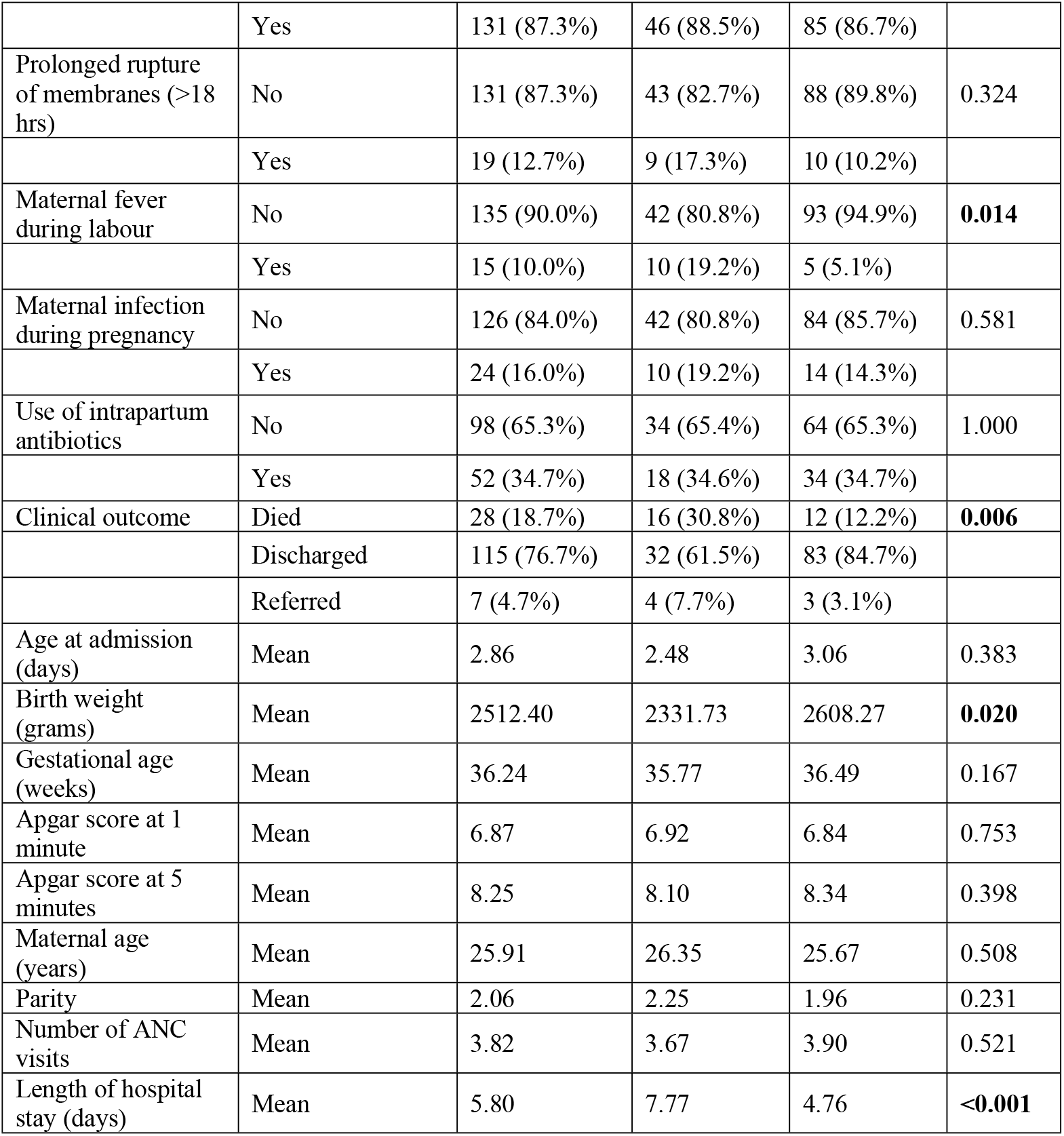
Socio-Demographic, Maternal And Clinical Characteristics By Sepsis Status.

### Factors associated with neonatal sepsis

In univariable analysis, increasing birth weight was associated with lower odds of neonatal sepsis (OR=0.947 per 100 g increase; 95% CI: 0.902-0.994; p=0.028), while maternal fever during labour was associated with higher odds. In the adjusted model, maternal fever during labour remained the strongest independent predictor (aOR=6.822; 95% CI: 2.032-22.905; p=0.002). Caesarean delivery was independently associated with sepsis (aOR=2.245; 95% CI: 1.050-4.801; p=0.037), while birth weight remained protective (aOR=0.891 per 100 g increase; 95% CI: 0.817-0.973; p=0.010). PROM showed increased odds but did not reach statistical significance (p=0.081).

**Table 2.** Univariable logistic regression: factors associated with neonatal sepsis.

| Variable | Comparison | Crude OR | 95%CI | P-Value |
| --- | --- | --- | --- | --- |
| Age at admission (days) | Per one-day increase | 0.963 | 0.882-1.052 | 0.405 |
| Sex | Male vs Female | 1.359 | 0.676-2.731 | 0.390 |
| Birth weight (per 100 g) | Per 100 g increase | 0.947 | 0.902-0.994 | 0.028 |
| Gestational age (weeks) | Per one-week increase | 0.926 | 0.830-1.034 | 0.173 |
| Mode of delivery | Caesarean vs SVD | 1.667 | 0.842-3.300 | 0.143 |
| Apgar score at 1 minute | Per one-point increase | 1.034 | 0.838-1.276 | 0.755 |
| Apgar score at 5 minutes | Per one-point increase | 0.914 | 0.746-1.121 | 0.390 |
| Maternal age (years) | Per one-year increase | 1.019 | 0.963-1.079 | 0.509 |
| Parity | Per one-birth increase | 1.154 | 0.910-1.463 | 0.236 |
| Antenatal care attendance | Yes vs No | 1.173 | 0.418-3.290 | 0.762 |
| Number of ANC visits | Per additional visit | 0.951 | 0.810-1.115 | 0.534 |
| PROM >18 hours | Yes vs No | 1.842 | 0.697-4.866 | 0.218 |
| Maternal fever during labour | Yes vs No | 4.429 | 1.425-13.760 | 0.010 |
| Maternal infection during pregnancy | Yes vs No | 1.429 | 0.585-3.486 | 0.433 |

**Table 3.**
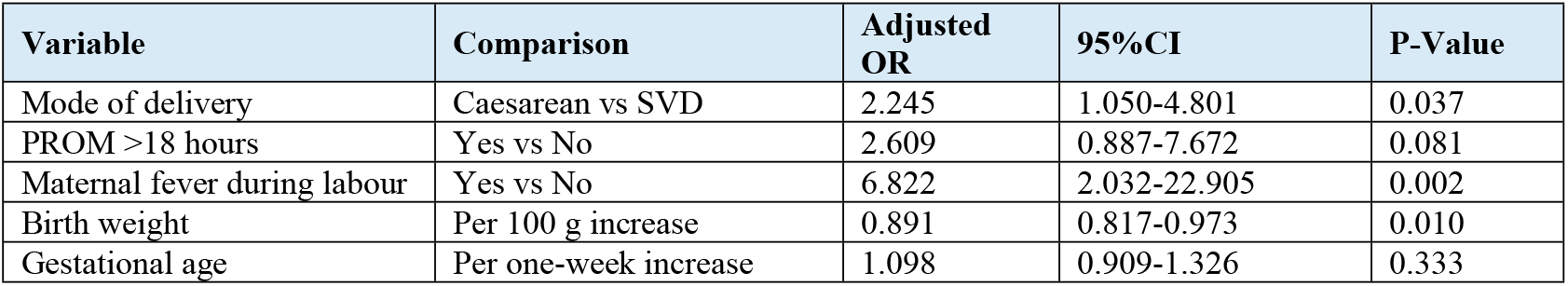
Multivariable logistic regression: independent factors associated with neonatal sepsis.

### Culture positivity

Among the 52 neonates classified as having sepsis, 20 had positive blood cultures, corresponding to a culture positivity rate of 38.5% (95% CI: 26.5%–52.0%). These 20 culture-positive cases represented 13.3% (95% CI: 8.8%–19.7%) of the full sample of 150 neonates. Subsequent bacterial-aetiology findings therefore use n=20 as the denominator, while the reported antimicrobial susceptibility summary uses n=19 because susceptibility results were available for 19 isolates.

### Bacterial pathogens isolated

Among culture-positive isolates, Gram-negative organisms predominated. *Escherichia coli* accounted for 25.0% of positive cultures, followed by *Klebsiella pneumoniae* (20.0%) and *Staphylococcus aureus* (20.0%). *Acinetobacter baumannii* and *Pseudomonas aeruginosa* each accounted for 10.0%.

**Table 4.** Bacterial pathogens isolated from neonates in the NICU.

| Identified organism | Gram stain | Frequency | Percentage of positive cultures |
| --- | --- | --- | --- |
| *Escherichia coli* | Gram-negative | 5 | 25.0% |
| *Klebsiella pneumoniae* | Gram-negative | 4 | 20.0% |
| *Staphylococcus aureus* | Gram-positive | 4 | 20.0% |
| *Acinetobacter baumannii* | Gram-negative | 2 | 10.0% |
| *Pseudomonas aeruginosa* | Gram-negative | 2 | 10.0% |
| *Enterococcus faecalis* | Gram-positive | 1 | 5.0% |
| *Coagulase-negative Staphylococcus* | Gram-positive | 1 | 5.0% |
| *Group B Streptococcus* | Gram-positive | 1 | 5.0% |

### Antimicrobial resistance patterns

Resistance was highest to ampicillin (78.9%), followed by gentamicin and ceftriaxone (52.6% each). Ciprofloxacin retained high activity, with 89.5% susceptibility, while all tested isolates were susceptible to meropenem.

**Table 5.**
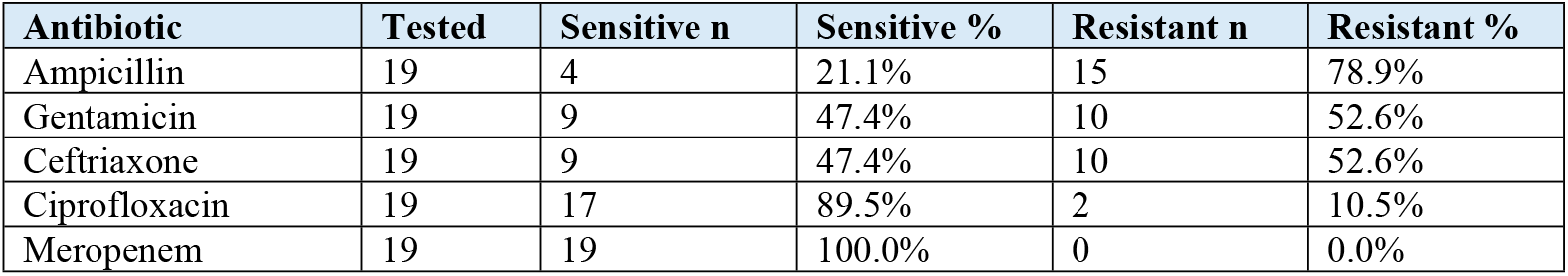
Antimicrobial resistance patterns among tested isolates.

| Antibiotic | Tested | Sensitive n | Sensitive % | Resistant n | Resistant % |
| --- | --- | --- | --- | --- | --- |
| Ampicillin | 19 | 4 | 21.1% | 15 | 78.9% |
| Gentamicin | 19 | 9 | 47.4% | 10 | 52.6% |
| Ceftriaxone | 19 | 9 | 47.4% | 10 | 52.6% |
| Ciprofloxacin | 19 | 17 | 89.5% | 2 | 10.5% |
| Meropenem | 19 | 19 | 100.0% | 0 | 0.0% |

## Discussion

The neonatal sepsis prevalence of 34.7% (95% CI: 27.5%–42.6%) at LUTH was close to the pooled prevalence reported in sub-Saharan African hospital-based studies, but higher than some facility estimates from Zambia and other settings [8,9]. Differences are likely to reflect referral patterns, case severity, diagnostic criteria, prior antibiotic exposure, laboratory capacity and infection-prevention conditions. As a tertiary referral centre, LUTH receives complicated neonatal cases, which may increase the observed burden relative to lower-level facilities. Importantly, only 20 of the 52 sepsis cases were culture-positive, giving a microbiological yield of 38.5% (95% CI: 26.5%–52.0%). Clinical sepsis and culture-confirmed bloodstream infection should therefore not be treated as interchangeable outcomes. Culture yield can be reduced by prior antimicrobial exposure, small neonatal blood volumes, intermittent bacteraemia and laboratory constraints.

Low birth weight was significantly associated with neonatal sepsis and remained protective when modelled continuously, with each 100 g increase reducing the odds of sepsis by approximately 11%. This is biologically plausible because low-birth-weight neonates have immature immune function, impaired barrier defenses and frequently require prolonged hospitalization and invasive care [3,5]. Maternal fever during labour was the strongest independent predictor, increasing the odds of neonatal sepsis almost sevenfold. Intrapartum fever may indicate intra-amniotic infection and increase the risk of vertical bacterial transmission [10]. PROM showed a positive but statistically non-significant association, possibly reflecting limited power for this exposure.

Caesarean delivery was independently associated with neonatal sepsis after adjustment. This association may reflect the clinical context in which caesarean sections are performed at a tertiary referral hospital, including obstetric emergencies, fetal distress and complicated pregnancies, rather than a direct causal effect of the procedure. Other factors including sex, gestational age, maternal age, parity, antenatal attendance and intrapartum antibiotic use were not independently associated with sepsis in this dataset.

Gram-negative bacteria predominated among culture-positive cases. Escherichia coli was the most frequent isolate, followed by Klebsiella pneumoniae and Staphylococcus aureus. This pattern is consistent with evidence from sub-Saharan Africa and Zambia showing an important role for Gram-negative organisms in neonatal bloodstream infection [5,6]. The presence of Acinetobacter baumannii and Pseudomonas aeruginosa, organisms frequently associated with healthcare environments, reinforces the importance of rigorous infection prevention and control within neonatal units.

The antimicrobial susceptibility findings raise concern about the continued effectiveness of commonly used empirical antibiotics. Resistance to ampicillin was 78.9%, while resistance to gentamicin and ceftriaxone was 52.6% for each agent. These findings are broadly consistent with prior Zambian and regional evidence of high resistance to first-line agents [5,6]. By contrast, susceptibility remained high to ciprofloxacin and complete to meropenem. Although this supports the retained activity of meropenem, it is a reserve antibiotic and should be protected through antimicrobial stewardship and culture-guided use to reduce selection for carbapenem resistance.

## Limitations

The study had several limitations. First, the retrospective design relied on routinely collected hospital records, creating a risk of information bias from incomplete, inconsistently documented or misclassified clinical variables. Missing data were not imputed, and estimates for incompletely documented variables therefore reflect available records. Second, the single-centre tertiary-hospital setting limits generalisability to district hospitals, primary-care facilities and neonates managed outside the NICU. Third, selection bias is possible because microbiological testing was not uniformly available for all suspected sepsis cases; more severely ill or more thoroughly investigated neonates may have been over-represented. Fourth, only 20 culture-positive isolates were identified and susceptibility data were reported for 19 isolates, producing imprecise organism-specific resistance estimates and limiting detailed stratification by early-versus late-onset sepsis. Finally, the study could identify associations but cannot establish causality because of its retrospective cross-sectional design.

## Conclusions

Neonatal sepsis at LUTH represents an important clinical and antimicrobial-stewardship challenge. The combination of substantial sepsis burden, incomplete culture confirmation and high resistance to commonly used first-line agents indicates that empirical treatment protocols should be periodically reviewed against current local microbiological evidence. Strengthening routine culture and susceptibility testing, developing a regularly updated facility-specific antibiogram, and integrating these data into infection-prevention and antimicrobial-stewardship programmes are important for improving treatment decisions and preserving effective antibiotics. These findings can inform institutional treatment guidance, while broader policy decisions require confirmation through larger prospective and multicentre surveillance studies.

## Data Availability

There are no legal or ethical restrictions.

## Recommendations

LUTH should strengthen infection prevention and control in the NICU, including hand hygiene, environmental cleaning and equipment decontamination. Blood culture and antimicrobial susceptibility testing should be obtained for suspected neonatal sepsis before antibiotic initiation where clinically feasible, and empirical treatment guidance should be regularly reviewed against local susceptibility data. Larger prospective multicentre studies and molecular epidemiological studies are recommended to improve generalisability and characterize resistance mechanisms and transmission pathways.

### List of abbreviations

AMR: Antimicrobial Resistance
AST: Antimicrobial Susceptibility Testing
CI: Confidence Interval
LMICs: Low- and Middle-Income Countries
LUTH: Livingstone University Teaching Hospital
NICU: Neonatal Intensive Care Unit
OR: Odds Ratio
PROM: Prolonged Rupture of Membranes
SVD: Spontaneous Vaginal Delivery

## Declarations

### Ethics approval and consent to participate

Ethical approval was obtained from the Mulungushi University School of Medicine and Health Sciences Research Ethics Committee and the National Health Research Authority of Zambia. Permission to use hospital records was obtained as required. The requirement for individual informed consent was waived because anonymized routinely collected hospital data were analysed.

### Consent for publication

Not applicable.

### Availability of data and materials

Data availability is subject to the ethical and institutional requirements governing the hospital records used in this study.

## Author contributions

LS and CS conceptualized the study, conducted the research, analysed and interpreted the data, and drafted the manuscript.

NT was involved in editing and writing of manuscript.

## Acknowledgements

The author acknowledges the supervisors, the School of Medicine and Health Sciences at Mulungushi University, and the management and staff of Livingstone University Teaching Hospital, particularly the NICU and laboratory personnel, for their support.

